# iGWAS: image-based genome-wide association of self-supervised deep phenotyping of human medical images

**DOI:** 10.1101/2022.05.26.22275626

**Authors:** Ziqian Xie, Tao Zhang, Sangbae Kim, Jiaxiong Lu, Wanheng Zhang, Cheng-Hui Lin, Man-Ru Wu, Alexander Davis, Roomasa Channa, Luca Giancardo, Han Chen, Sui Wang, Rui Chen, Degui Zhi

**Affiliations:** Department of Molecular and Human Genetics, Baylor College of Medicine, Houston, Texas 77030, USA; School of Biomedical Informatics, The University of Texas Health Science Center at Houston, Houston, Texas 77030, USA; School of Public Health, The University of Texas Health Science Center at Houston, Houston, Texas 77030, USA; Department of Ophthalmology, Stanford University School of Medicine, Stanford, CA 94305, USA; Department of Ophthalmology and Visual Sciences, University of Wisconsin, Madison, Wisconsin 53726; Human Genetics Center, Department of Epidemiology, Human Genetics and Environmental Sciences, School of Public Health, The University of Texas Health Science Center at Houston, Houston, Texas 77030, USA

**Keywords:** Deep Learning, Artificial Intelligence, Retina image, Imaging genetics, GWAS

## Abstract

Existing imaging genetics studies have been mostly limited in scope by using imaging-derived phenotypes defined by human experts. Here, leveraging new breakthroughs in self-supervised deep representation learning, we propose a new approach, image-based genome-wide association study (iGWAS), for identifying genetic factors associated with phenotypes discovered from medical images using contrastive learning. Using retinal fundus photos, our model extracts a 128-dimensional vector representing features of the retina as phenotypes. After training the model on 40,000 images from the EyePACS dataset, we generated phenotypes from 130,329 images of 65,629 British White participants in the UK Biobank. We conducted GWAS on three sets of phenotypes: raw image phenotype, phenotypes derived from the original photos; retina color, the average color of the center region of the retinal fundus photos; and vessel-enriched phenotypes, phenotypes derived from vasculature-segmented images. GWAS of raw image phenotypes identified 14 loci with genome-wide significance (p<5×10^-8^ and intersection of hits from left and right eyes), while GWAS of retina colors identified 34 loci, 7 are overlapping with GWAS of raw image phenotype. Finally, a GWAS of vessel-enriched phenotypes identified 34 loci. While 25 are overlapping with the raw image loci and color loci, 9 are unique to vessel-enriched GWAS. We found that vessel-enriched GWAS not only retains most of the loci from raw image GWAS but also discovers new loci related to vessel development. Our results establish the feasibility of this new framework of genomic study based on self-supervised phenotyping of medical images.

## Introduction

Although genome-wide association studies (GWAS) have successfully identified thousands of genetic associations, most existing GWAS are based on a set of predefined phenotypes. While these phenotypes encode valuable biomedical knowledge, they are also biased by current clinical practice and epidemiological studies. In addition, as the granularity of phenotype code is often limited, it is often not sufficient to capture the complexity of human physiology and pathology in their entirety. Therefore, deriving new phenotypes beyond expert curation would enable the discovery of new genetic associations.

Medical imaging is a rich resource for phenotype discovery. Through rapid technological advancements, modern medical imaging offers unprecedented details about a patient’s physiological condition and can be a high-content phenotyping modality. Existing imaging GWASs have leveraged imaging-derived phenotypes (IDPs)^1–3^. These IDPs were typically designed by imaging experts and generated by special-purpose image processing pipelines. Recently, machine learning, especially supervised deep learning (DL), is used to automatically generate IDPs^4–6^. These methods were trained by learning from data labeled by experts and identified new loci in GWAS^2, 7, 8^. However, although supervised DL can vastly improve the efficiency of image labeling, it fails to provide phenotypes beyond those defined by experts. In addition, although these phenotypes are derived for medical practice, clinical decision processes, and natural-language-based reporting, they often do not comprehensively capture the imaging content. There are limitations to the amount of information a human eye can extract from images. Many meaningful imaging features, some of which might be used implicitly by physicians, may not be verbalized in medical reports. In addition, there may be physiologically informative features that are present in the image but are completely missed or ignored by readers. For example, Google’s DL algorithm extracted novel features from retinal images, such as age, gender, and smoking status, that are not readily apparent to expert human graders^9^. Following studies identified features such as refractive error and anemia from retinal images^10, 11^. These results suggest that additional information beyond human curation may be encoded within imaging data, and new methods are needed to extract such information.

Here, we have designed a new framework of genome-wide genotype-phenotype association study by performing unsupervised image-based genome-wide association studies (iGWAS). For phenotype discovery, instead of supervised learning that relies on labels from expert annotations, unsupervised deep learning is applied to an image to capture its intrinsic contents^12–15^. Endophenotypes generated by the deep learning model are then subjected to GWAS to identify associated genomic loci.

We tested this new approach using human fundus images. First, we derived endophenotypes from the raw color fundus images, which likely capture the overall content of the image. In addition, we derive endophenotype from vessel-segmentation of the raw images, which likely capture features enriched for the vascular structure of the retina. Vessel-enriched features are chosen because retinal vasculature of healthy individuals remains relatively stable since development, making it a suitable phenotype for genetic association studies. In addition, defects in the blood vessels in the retina lead to many human retinal diseases, such as diabetic retinopathy, and the feasibility of learning retinal vasculature representations as biomarkers has already been investigated^16, 17^. We constructed a contrastive loss function over an Inception V3 architecture to learn a representation that captures the intrinsic retinal features of individuals. Our neural network outputs 128 endophenotypes representing the input image, either raw color image or vessel-segmented image. After training on 40,000 images from EyePACS, our model generated phenotypes from 130,329 images of 65,629 British White participants in the UK Biobank. We conducted three sets of GWAS analyses: the raw fundus image endophenotypes, the vessel-enriched endophenotypes, and the average colors of the central patch representing retina colors. The retina colors were included in our analyses to aid the interpretation of our results as they are the most prominent feature of the fundus image. In addition, a follow up functional assay verified the role of the *WNT7B* gene, a novel candidate locus, in retinal vascular development *in vivo*.

## RESULTS

### Overall iGWAS framework

The core component of iGWAS is a phenotyping (encoder) neural network that generates endophenotypes, which are in turn associated with genotypes by GWAS (an example of iGWAS for retinal images is shown in **Figure 1**). Distinct from traditional phenotypes labeled by experts or by AI trained via supervised learning, iGWAS’s encoder network is trained by self-supervised learning to discover new phenotypes. We thus named it as **S**elf-**Su**pervised **P**henotyp**er** (SSuPer). Popular self-supervised learning losses, such as contrastive losses^12, 14, 18^ and reconstruction losses^19^, are used to extract coherent and biologically relevant features of individuals. We used a contrastive loss to learn features that are consistent between the images from the same person. The resulting “embedding vector,” the output of the encoder, is treated as “endophenotypes” for downstream GWAS analysis.

**Figure 1:**
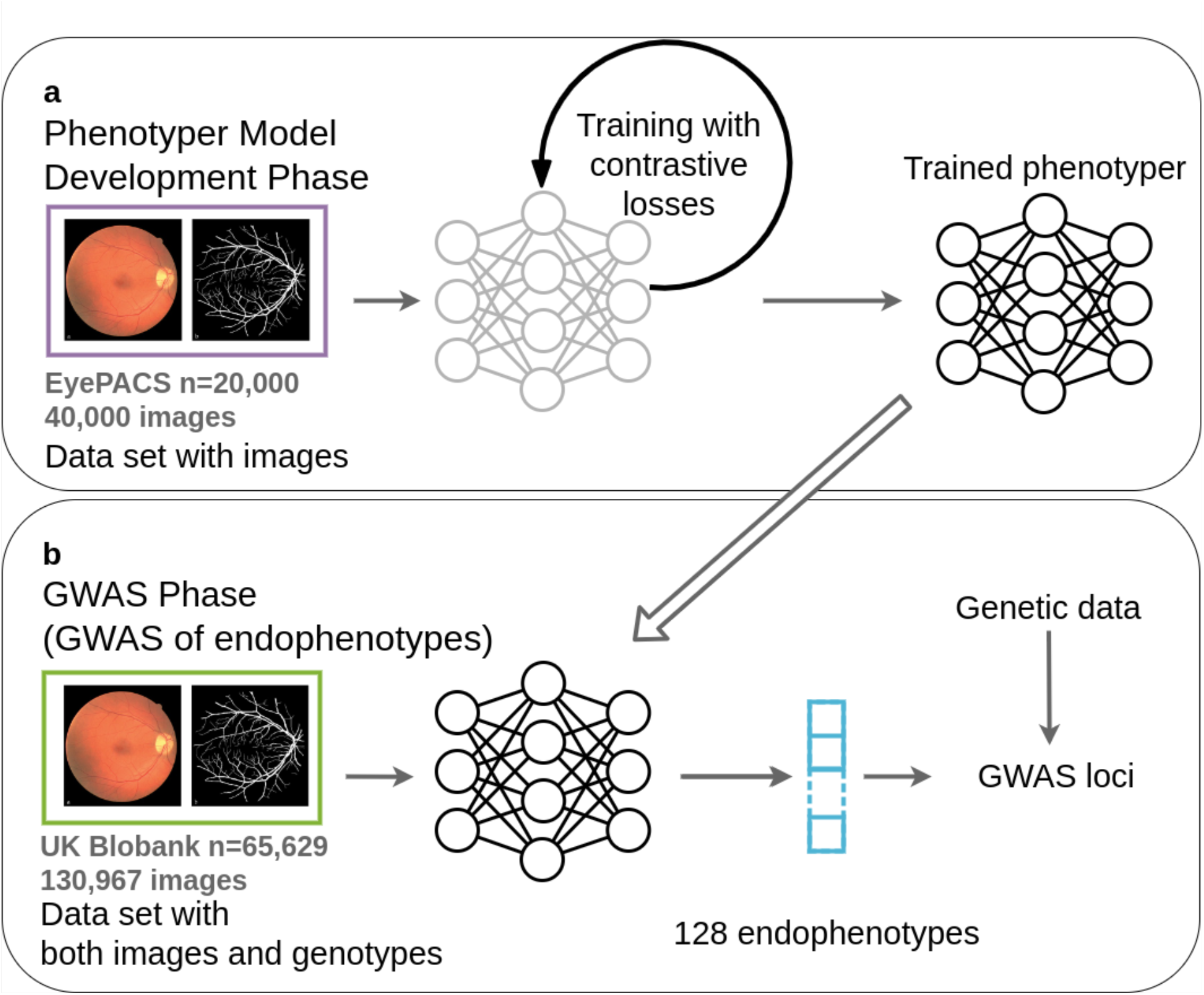
iGWAS of endophenotypes from retinal fundus images. (**a**) Using raw fundus images or segmented vessel mask images derived from fundus images (segmentation network omitted for conciseness) in EyePACS, we developed phenotyper neural networks that optimize contrastive losses; (**b**) Using the trained phenotypers, we generated 128 endophenotypes for each raw and vessel image in the UK Biobank vision cohort and do GWAS on these endophenotypes to identified independent loci.

The iGWAS approach is executed in two phases: the model development phase and the GWAS phase. In the model development phase, a “phenotype development set” is used to train the embedding network. The phenotype development set is a collection of images from individuals, whose genotype data are not needed. The result of the model development phase is a trained neural network model, SSuPer, that can transform an input image into a set of self-supervised image-derived phenotypes (SS-IDPs). In the GWAS phase, the trained SSuPer from the model development phase is used to generate SS-IDPs for images from the “GWAS set,” a dataset containing both images and genotypes of a different cohort of individuals. The SS-IDPs are then tested for association with genome-wide markers.

### Overall data analysis strategy for generating raw-image and vessel-enriched endophenotypes from fundus images

In this study, we designed and implemented the iGWAS approach to encode retinal features from fundus images. For the phenotype development set, we used data from EyePACS, a large public collection of 88,702 fundus images (see **Methods: dataset extraction**). After quality control (see **Methods: Image quality control**), 40,000 top quality images were used (**Supplementary Table 1**). For the GWAS set, we used fundus images and genotype data of 65,629 British White UK Biobank participants. Although the demographics of the EyePACS and UK Biobank cohorts do not match exactly, we reasoned that some characteristics of their fundus images should be similar, so we expect the features learned from EyePACS can be generalized to UK Biobank.

First, the EyePACS fundus images are directly fed into the encoder neural network to generate raw image endophenotypes. A convolutional neural network (CNN) based on the Inception^20^ architecture is used because it is proven to deliver good results for modeling images.

In parallel, the EyePACS fundus images were also fed into a vasculature segmentation network to generate vessel masks, which were then treated as the inputs to an embedding network to generate the vessel-enriched endophenotypes. We expected that using the segmentation mask would filter out much of the other information in the raw input image and thus result in images with enriched vasculature information, so that the endophenotypes would mainly be related to the vasculature as well.

We found some of the endophenotypes strongly correlated with the color in both raw image derived endophenotypes and the vessel segmentation mask derived endophenotypes. Therefore, to account for the “retinal color,” defined as the average intensities of the red, blue, and green channels of the central patch of the fundus image, were considered as additional phenotypes in subsequent analysis (see Methods). While the definition of retinal color may not fully account for change in illumination, texture of the retinal pigment epithelium, retinal lesions, and optic disk, it captures coarse-grained information of the retinal background. We conducted GWAS analyses for the three sets of phenotypes: 128 raw image endophenotypes, 128 vessel-enriched endophenotypes, and 3 retina colors (RGB channels). To aid in interpretation of the endophenotypes, we conducted univariate and correlation analyses among endophenotypes and between endophenotypes and relevant eye phenotypes. The overall pipeline is shown in **Supplementary Figure 1**.

### Design of encoder network that captures coherent features of fundus images from the same person

To generate an embedding vector that represents the inherent biological features of an individual, we leverage a self-supervised metric learning approach that was described in ArcFace^21^, a widely adopted algorithm that is used to extract features for developing human face recognition methods, with some technical modifications detailed in methods (see **Methods: Embedding neural network**). Inception v3, which has been demonstrated to be capable of capturing complex information within fundus images, was used as a backbone architecture for the metric learning^22^. The output of our embedding network was designed to be a 128-dimensional vector, based on previous work showing that 128-dimensional vectors are sufficient to represent complex datasets^23, 24^. Our ArcFace loss function is a contrastive loss that first projects the embedding vector to the unit sphere and then optimizes the contrast between the embeddings from the eyes of the same person and the embeddings from different people by minimizing the angular distance between the embeddings of left and right retinas from the same individual while keeping the embeddings from different individuals at least some margins apart (**Figure 3**). We reasoned that if the trained model manages to capture real biologically relevant features, embeddings between an individual’s left and right fundus images should be more similar than those from different individuals, previous work also showed that genetic relatedness can be estimated from pairs of fundus images^25^. Details of model design and training are described in the **Methods: Embedding neural network**.

For the vessel segmentation network, we chose a patch-based vessel segmentation network with the classic U-net architecture^26^ (See **Methods: Fundus image segmentation**). This is an easy choice because vessel segmentation of fundus images is a well-studied problem with mature methods^27–29^.

### Training of encoder networks

For 88,724 images from EyePACS, 54,992 passed our quality filter network (quality score > 0.5) (see Methods). 40,000 top quality images (quality score > 0.95) were selected as we reasoned that this balance point of sample size and sample quality is sufficient for training the main SSuPer network. The characteristics of the EyePACS dataset are shown in **Supplementary Table 1**.

Both the segmentation and the embedding networks were trained using standard gradient descent. See Methods for details. To verify the performance of the SSuPer embedding network, we compared the matched pairs (left and right eye of the same person) and random pairs (**Figure 2b**). As expected, there is a clear separation in the distribution of cosine distance between matched pairs and random pairs (see **Supplementary Table 2** for quantification). Although less than that of EyePACS, the separation of matched and random pairs was clearly observed in UK Biobank, indicating the segmentation and embedding models are transferable and indeed capture the intrinsic features of the fundus images. Therefore, we directly applied the embedding networks trained using EyePACS to the UK Biobank data without fine-tuning. Of note, we observed a cone effect that the cosine similarity between any pair of embeddings is almost always greater than 0, which is a general phenomenon for deep neural networks^30^. Interestingly, the cone effect is more prominent for the embeddings of vessel mask images, which may indicate that the contrastive learning task is more difficult for the vessel mask images.

**Figure 2:**
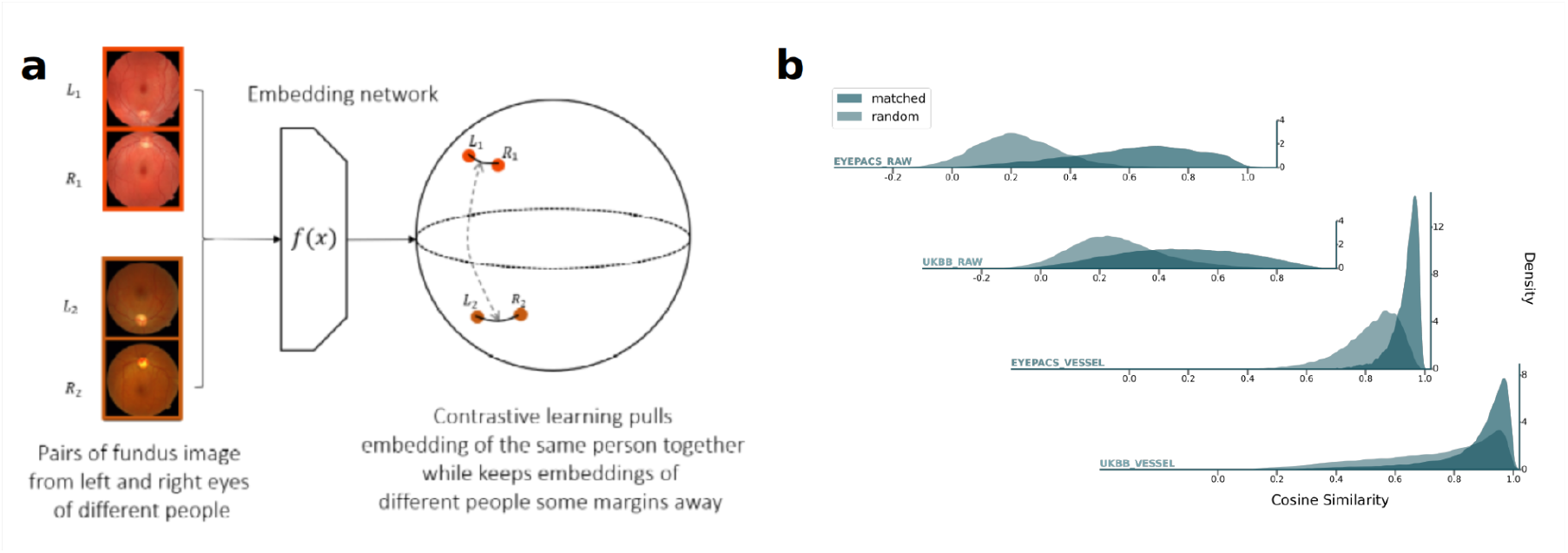
Contrastive loss for deriving phenotypes coherent across images from the same person. (a) Contrastive loss is designed to map images from the same person to be closer while keeping images from different persons apart. The trained endophenotype vectors for raw fundus image embedding and for vessel-enriched embedding (**b**) of the same persons reflect the design of contrastive learning in both the training set (EyePACS) and the test sets (UKBB). The distributions of the matched pairs (images from the same person) and the random pairs are separated. The distributions were estimated using Scott’s kernel with an additional multiplicative factor of 0.5 to smooth the curve.

### Descriptive analysis of endophenotypes in UK Biobank fundus photos

We conducted our analysis using 65,629 British White participants from the UK Biobank who had available fundus images (see **Methods: Dataset extraction**). For each participant, we chose the first image for each eye, resulting in 130,329 images. Basic demographic description of this dataset is shown in **Supplementary Table 3**. Retina colors were also extracted as phenotypes. The central patch of the image (the fovea region) was used because it has more pigment and of low vessel density, providing a cleaner estimate of the retinal color (see **Methods: Color GWAS**).

Univariate distributions of the endophenotypes generated by our embedding networks is shown in **Supplementary Figure 2**. Interestingly, we found that while most raw image endophenotypes have unimodal bell-shaped distributions, some vessel-enriched endophenotypes have bimodal or multimodal distributions. Meanwhile, examining their pairwise correlations showed that each endophenotype group has strong internal correlations (more so in vessel-enriched endophenotypes) but the two groups themselves are weakly correlated (**Figure 3**), indicating that these two embeddings capture different aspects from images.

**Figure 3.**
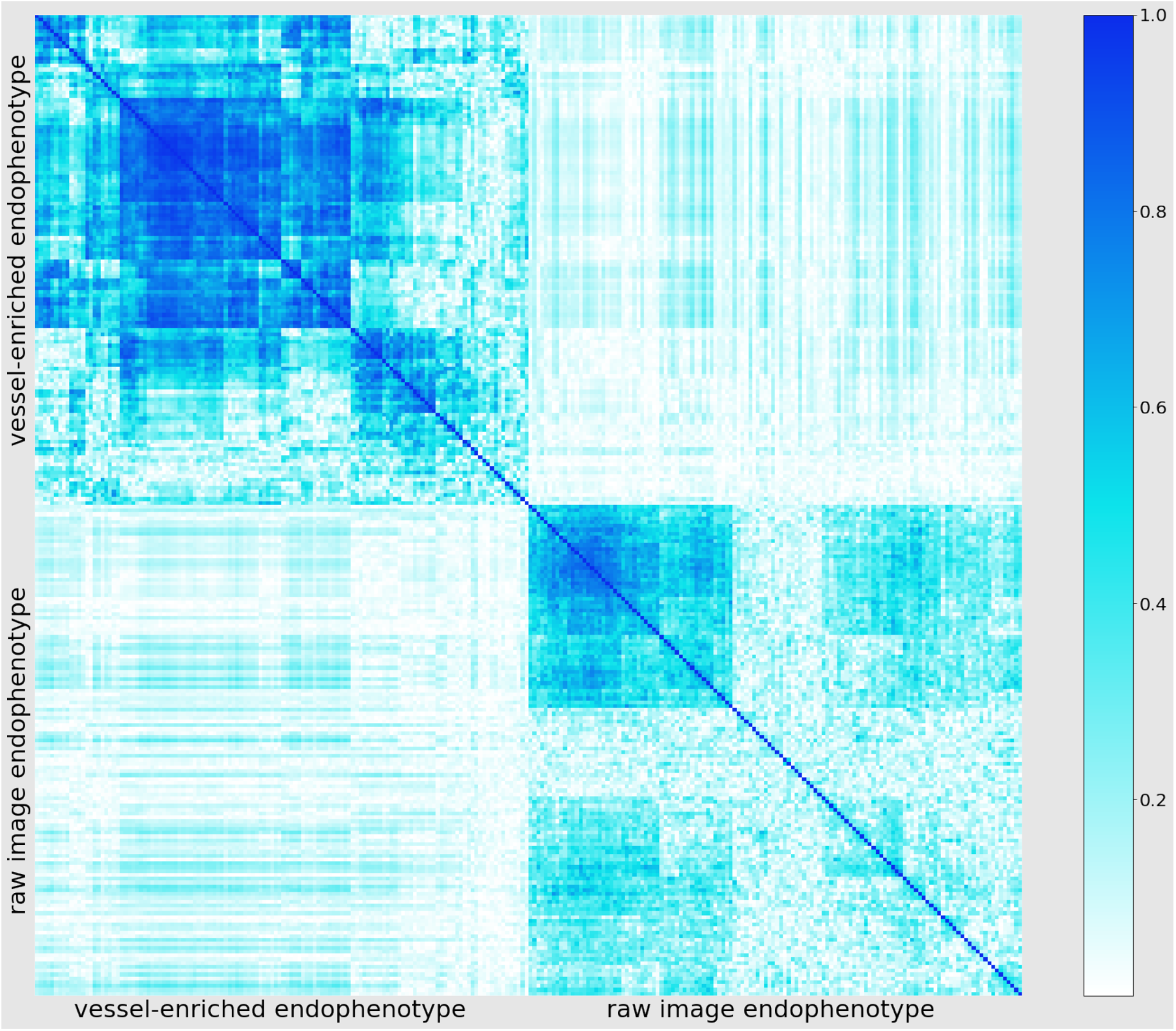
Absolute correlations among 128 vessel-enriched (upper left) and 128 raw image-derived (lower right) endophenotypes. While some correlations within each endophenotype block are observed by hierarchical clustering, there is a lack of correlations across vessel-enriched and raw-image derived endophenotypes.

### iGWAS: GWAS of endophenotypes

To identify genetic factors associated with endophenotypes, GWAS was performed for each of the 128 dimensions from all 130,329 images using linear mixed models as implemented by BOLT-LMM^31^, adjusted by age, sex, and ancestral principal components (PCs) (see **Method: Endophenotype GWAS**). Analyses were conducted separately for the left and right retinal images. Their results were not meta-analyzed because the endophenotypes of the two eyes may be correlated due to training. Instead, we pooled the results from the two eyes and took the intersection of the significant hits, and only the more significant p-value between the two eyes was reported. Since the endophenotypes were derived without the direct use of any genetic information, we expected there to be minimal genomic inflation for the GWAS. Indeed, we observed that the genomic inflation factor was well-controlled (λ_GC_≈1) (**Supplementary Figure 3**), though some endophenotypes had slightly higher (1.099) inflation factors, indicating potential polygenic genetic architecture.

For raw image endophenotype GWAS, we identified 2,150 SNP-endophenotype pairwise association signals from 113 SNPs (**Supplementary Table 4**) showing genome-wide significance (p-value<5×10^-8^) (**Figure 4**). These SNPs were clustered into 14 independent loci (**Table 1**) (see **Methods: Endophenotype GWAS**).

**Figure 4:**
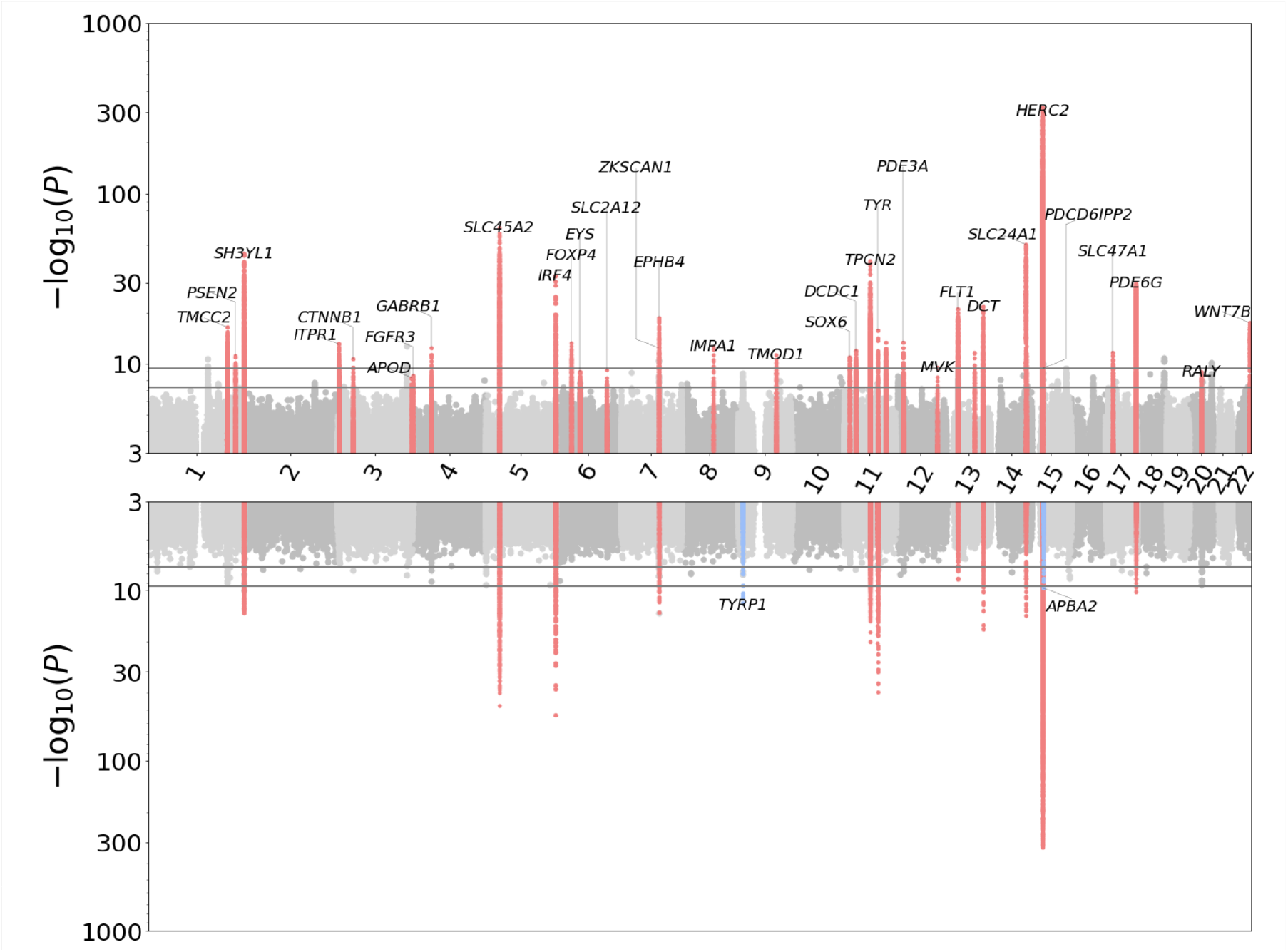
Aggregated Miami plot with Manhattan plots of 128 vessel-enriched endophenotypes on the top, and that of raw image endophenotypes on the bottom. The two horizontal lines indicate significance levels set for individual GWAS (p=5 × 10^-8^) and all phenotypes (p=5 × 10^-8^/128). The red peaks are the vessel-enriched endophenotype associated loci that satisfy selection criteria defined in **Methods: Endophenotype GWAS**. The blue peaks are unique to raw image endophenotype GWAS.

**Table 1:**
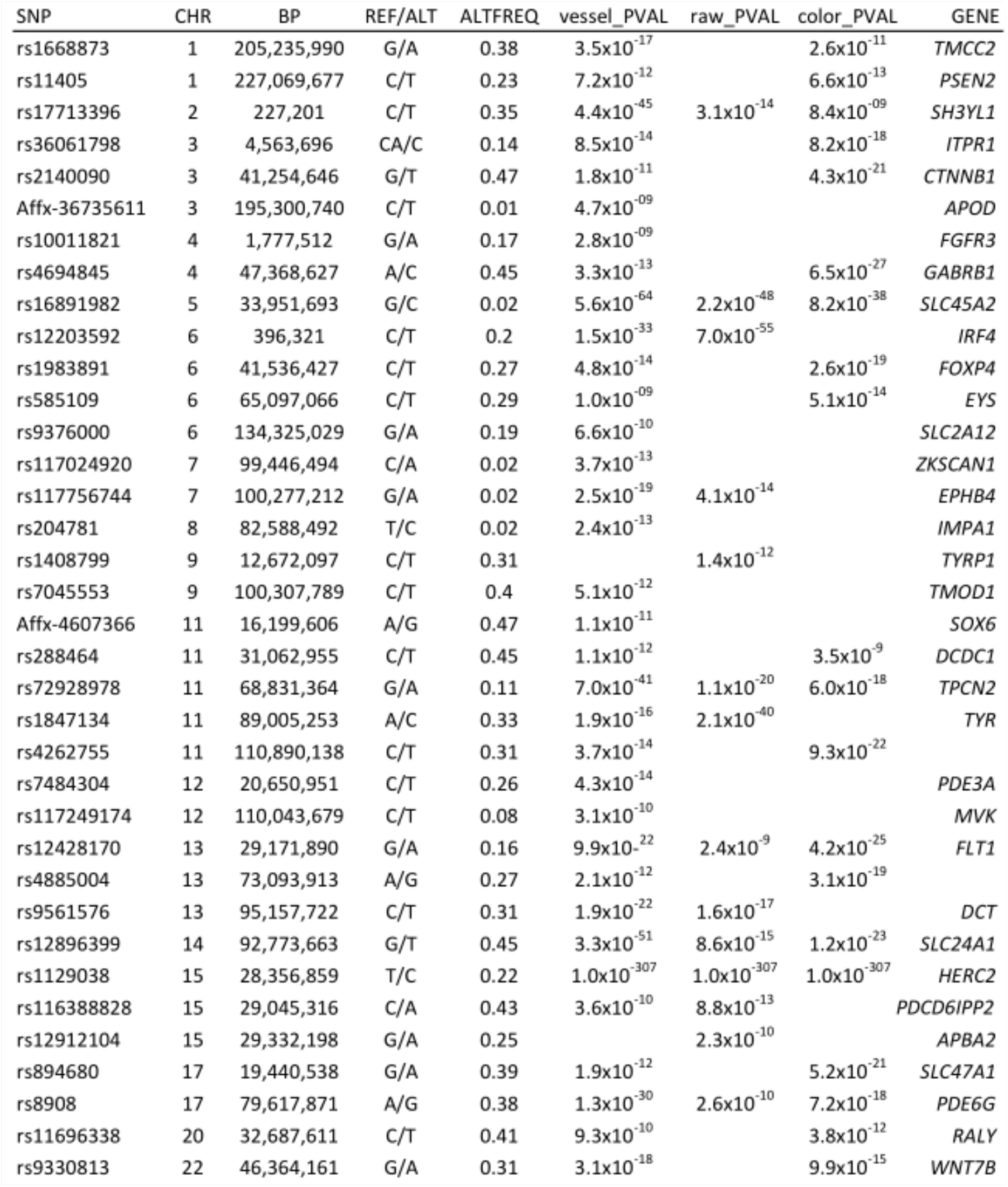
Loci significantly associated with raw-image or vessel-enriched endophenotypes found by iGWAS. BP is in GRCh37 coordinate. The vessel_PVAL and raw_PVAL columns contain the most significant p-value (min P) of the vessel-enriched endophenotype and the raw image endophenotype GWAS among all endophenotypes at each locus. Non-significant p-values are omitted. Candidate genes for each locus were annotated based on their distance from the leading SNPs and annotated function.

The mean and standard deviation of the heritability from LD score regression (**Supplementary Figure 4, Supplementary Table 10**) of raw image endophenotype are 0.04 and 0.05 while those of vessel-enriched endophenotypes are 0.10 and 0.05 (T test p-value = 1.8×10^-37^).

For vessel-enriched endophenotypes, we identified 4,986 association signals from 176 SNPs (**Supplementary Table 5**) showing genome-wide significance (p-value<5×10^-8^) (**Figure 4**). These SNPs were merged into 34 independent loci (**Table 1**) (see **Methods: Endophenotype GWAS**). Additionally, to test if the vessel-enriched endophenotypes can be represented by lower numbers of components and to evaluate the potential of removing variations in unwanted directions, we did principal component analysis for the vessel-enriched endophenotypes, selected the top 99% variance explaining components for GWAS and calculated the heritability (see Methods: PC GWAS and heritability calculation and **Supplementary Table 11, 12**). We found that the top variance explaining components have lower heritabilities than many vessel-enriched endophenotypes and the heritability decays very fast in these components, indicating that the principal components are not effective at capturing genetic variations.

For vessel-enriched endophenotype GWAS, we make the following observations. Very recently, in a GWAS of retinal vessel tortuosity using fundus images^32^, tortuosity was determined by a non-deep learning software. Interestingly, two of the 34 of loci identified in our study overlapped with the top 10 SNPs shown in the tortuosity study^32^, at chr12:*PDE3A* and chr15:*HERC2/OCA2*. Furthermore, at least five candidate genes we identified, including *FLT1*, *EPHB4*, *WNT7B*, *SOX6*, and *APOD*, have been reported to have vessel-related functions. For example, highly significant SNPs (p-value = 4×10^-23^) were found around *FLT1*, which is a negative regulator of *VEGF* and is known to play an important role in retinal vessel development^33^. Similarly, a strong association was found with erythropoietin-producing hepatocellular receptor B4 (*EPHB4*) (**Supplementary Figure 5a**) , which is essential in vessel development^34^. Modulation of *Ephb4* activity in the mouse retina was found to alter retinal neovascularization^35, 36^. Another identified locus was centered at *WNT7B* (**Supplementary Figure 5b**), which is known to be important in blood brain barrier development^37, 38^. Similarly, *SOX6* has been implicated in arteriole development in kidney^39^ and *APOD* has been linked to chorioretinal blood vessel formation^40^. Interesting, these loci in *SOX6* and *APOD* have not been associated with retinal vasculature measurement phenotypes (EFO ID: EFO_0010554) in previous GWAS studies^41–43^ as documented in GWAS Catalog, further indicating that our framework is an effective, novel approach for identifying new genetic loci associated with phenotypes captured by imaging.

For raw image endophenotype GWAS, 2 additional loci were identified beyond what was found in the vessel-enriched endophenotype GWAS: *TYRP1* and *APBA2*. *TYRP1* and *APBA1* are both associated with eye color^44, 45^.

### GWAS of retina color

Pigmentation of the human body, such as hair, skin, and iris, is strongly influenced by genetics. As the color of the human retina is influenced by factors such as the level of pigmentation of retinal pigment epithelium (RPE) and choroid blood vessels, we tested if genomic loci associated with retina color can be identified through genome association study of fundus images (**Methods: Color GWAS**). While association of iris color has been conducted^46^, no direct association studies of retinal color using fundus images have been conducted. In our study, significant genome-wide association (p<5×10^-8^, and intersection between hits from fundus images of left and right) was obtained for a total of 175 SNPs (**Supplementary Table 6** and **Supplementary Figure 6**) from 34 independent loci (**Supplementary Table 7**).

We found 13 out of the 34 retina color loci overlapped with previously reported GWAS loci for “hair color”^47^, “eye color”^48^, and “skin pigmentation”^48^ in the GWAS catalog (**Supplementary Table 7**, **Supplementary Figure 8**, see **Supplementary Table 7** for details), supporting the validity our approach. Interestingly, many genes from unique loci identified in this study can be linked to pigmentation pathways (**Supplementary Table 7**). For example, mutations in *FGFR3* lead to familial acanthosis nigricans, which results in skin pigmentation abnormalities^49^. In addition to pigmentation, it is interesting to note that 6 of the 34 loci overlap with loci previously reported to be associated with macular thickness (**Supplementary Table 7**), including *DCDC1*, *TPCN2*, *NCAM1*, *HERC2*, *PDE6G*, and *WNT7B*.

### Genetic correlation analyses of endophenotypes

To further interpret these vessel-enriched endophenotypes, we correlated them with other traits that are related to retinal phenotypes. We conducted genetic correlation using summary statistics as they are easier to access and are suggested to be a good surrogate for phenotypic correlation^50, 51^. We included traits that either have GWAS hits near the endophenotype GWAS loci (within 200kb) or are known to be related to retinal or corneal disorders, and whose genetic summary statistics for UK Biobank data are available (see **Methods: Genetic correlation**). Corneal phenotypes (H15-H22 Disorders of sclera, cornea, iris and ciliary body and H18 Other disorders of cornea from GeneAtlas^52^) were included because they may affect refractive error, which can have a detectable effect on the fundus images. We found that many endophenotypes (both raw image derived and vessel-enriched) are genetically correlated with skin/hair pigmentation and retinal color after Bonferroni correction (corresponds to p-value threshold of 0.05/128). Other nominally significant genetically correlated pairs (not significant after Bonferroni correction) include correlations between both endophenotypes and cardiovascular disease, diabetes, lung function and blood pressure. Vessel-enriched endophenotypes additionally show genetic correlations with cornea disorder, glaucoma, arterial diseases and retinal disorder (**Supplementary Table 8, 9**).

We also correlated (phenotypically and genetically) both the raw image endophenotypes and vessel- enriched endophenotypes with fundus background color and found that the latter are more strongly correlated with fundus background color. The strong correlation between vessel-enriched endophenotypes and retinal color is possibly due to the vessel segmentation masks lacking texture information which is abundant in raw images, so the phenotyping network for the vessel segmentation masks has to rely more on the color information (**Supplementary Table 8, 9**).

### Functional validation of candidate gene involved in retinal vessel development

To validate the candidate genes identified from our vasculature GWAS, we sought to test if *Wnt7b* functions in retinal vessel development. Although mice with partial loss of function of *Wnt7b* mutation show persistence of the hyaloid vessel, involvement of *Wnt7b* in the retinal vasculature has not been reported^53^. Four shRNAs (shRNA80-83) targeting *Wnt7b* were designed and tested in cell culture for their knockdown efficiency. Greater than 70% knockdown efficiency was observed for all shRNAs with shRNA83 having the highest knockdown efficiency (**Figure 5a-f**). To test the function of *Wnt7b* in the retina, shRNA80 and shRNA83 were electroporated into mouse retinas *in vivo* at postnatal day 0 (P0) as described^54^ (**Figure 5g**). Thirty days after electroporation, the retinas were harvested following injection of NHS-biotin vessel tracer^55^. When *Wnt7b* was knocked down in the retina, the total vessel area was significantly increased in the intermediate vascular plexus and reduced in the deep vascular plexus (**Figure 5h-k**). Similarly, the vessel branch-points were significantly increased in the intermediate and superficial plexuses and decreased in the deep vascular plexus when *Wnt7b* was depleted from the developing retina (**Figure 5l-n**). These results demonstrate that *Wnt7b* plays an important role in normal retinal vascular development.

**Figure 5:**
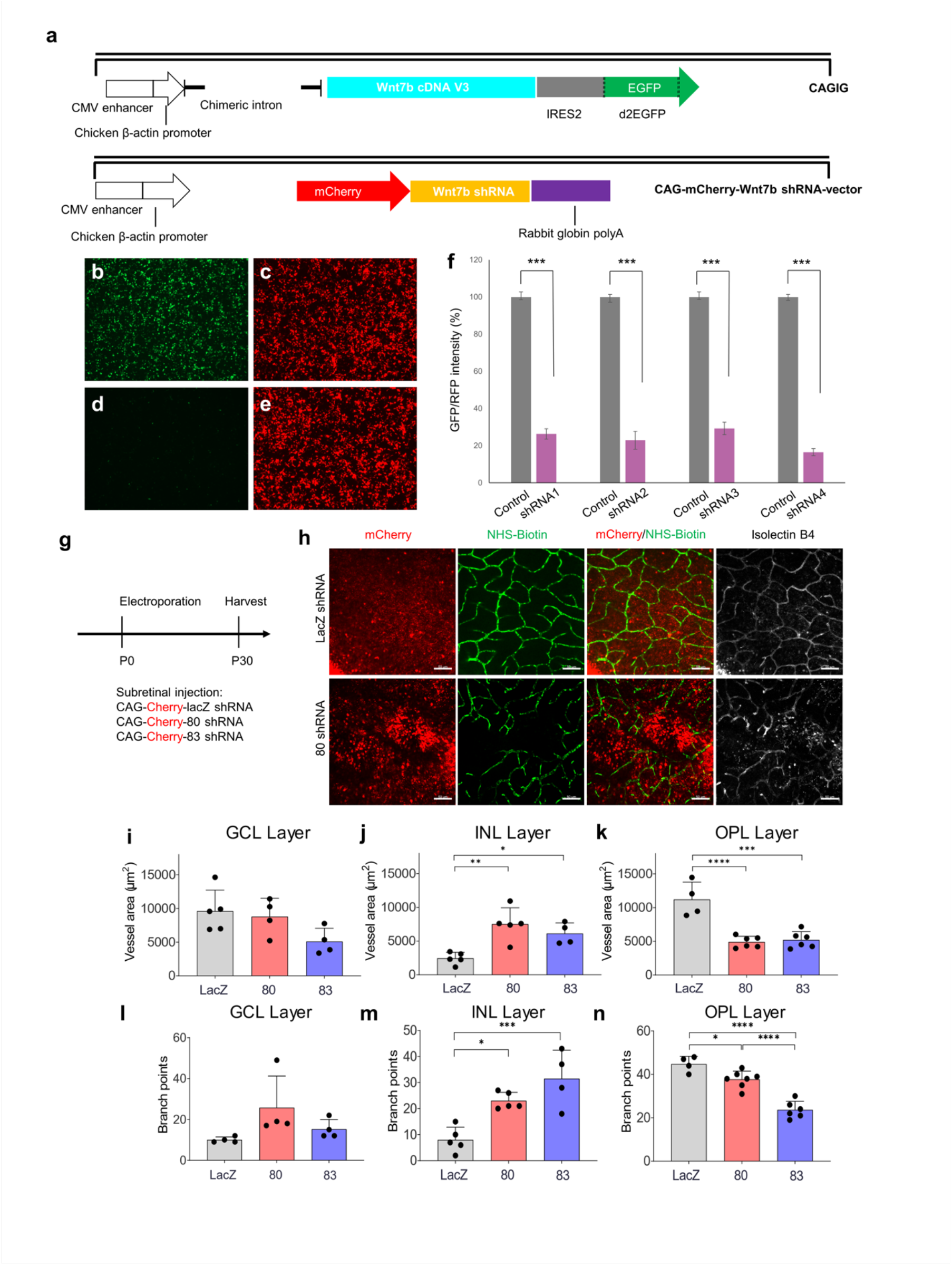
Depletion of *Wnt7b* in developing retina induced defects in vascular development. **(a)** Schematic constructs of *Wnt7b* cDNA- and shRNA-expressing vectors. **(b-e)** Figures demonstrating HEK293T cells under GFP and mCherry channels after 24-hours’ transfections. **b:** *Wnt7b* cDNA only. **c:** *Wnt7b* shRNA80 only. **d:** GFP channel of *Wnt7b* cDNA + shRNA80. **e:** mCherry channel of *Wnt7b* cDNA + shRNA80. **(f)** Knockdown efficiencies of each shRNA after 24 hours transfection was calculated. Gray column: The ratio of cDNA only GFP fluorescent intensity over shRNA only mCherry fluorescent intensity. Magenta column: the ratio of GFP over the mCherry fluorescent intensity after a combo of cDNA and shRNA transfections. Statistical differences were determined by Two-tailed T-test. *** represents a significant difference P-value <0.001. **(g).** The experimental design. **(h).** Knock down of *Wnt7b* (shRNA 80) led to abnormal vessels in the retina in the outer plexiform layer (OPL). NHS-Biotin: tracer for vessels. Isolectin B4: vascular cell marker. **(i-n).** Quantification of vessels in superficial, intermediate and deep vascular plexuses in the retina. Vessel area and branch points were quantified. Data are mean ± SEM (n = 3 in control group, six lobes of retina were included; n = 2 in shRNA 80 group, four lobes of retina were included; n = 2 in shRNA 83 group, four lobes of retina were included). Statistical differences were determined by one-way ANOVA with a post Tukey’s multiple comparisons test, comparing the shRNA 80 and 83 groups with the control group. *p < 0.05, **p < 0.01, *p < 0.001 and ****p < 0.0001. Scale bar: 50 μm.

## Discussion

Our work is one of the first proof-of-concept studies of a self-supervised learning-based phenotype discovery method for imaging GWAS. With no expert supervision, our method was able to extract endophenotypes and identify genes relevant to retinal vessel development, including a new locus that was validated experimentally.

While there have been previous imaging GWAS on DL-based phenotyping, they either used expert-defined phenotypes^7,43^ or clustering of dense representational vectors into subtypes. We directly use the dense vectors, which contain more information than the subtype cluster labels, as phenotypes. There are a handful of studies that use final or intermediate layers of the neural network as phenotypes, but these networks were trained in a supervised fashion using external labels (e.g., age^56^ or eye diseases^57^) or via transfer learning^58^. Of note, there is another contrastive learning approach, ContIG, for phenotyping the retina fundus images by maximizing cross-modality matching between the image part and the genetic part of the same individuals^59^. However, we found that the GWAS of ContIG derived phenotypes returns less loci than what derived by iGWAS, may be due to the fact that part of the sample sizes was consumed by the model training and thus GWAS sample size is limited. On the other hand, iGWAS does not require data sets with both images and genetic data to train the encoder, and may have a wider range of applicability.

Our iGWAS framework is flexible and can be adapted and extended in various ways. To study retinal vasculature, our current approach includes a segmentation step that generates vessel masks, and the embeddings are subsequently derived from these predicted vessel masks. To capture additional information in fundus images, such as the morphology of the optic disc, hemorrhages, exudates, or the pigmentation level, alternative preprocessing/segmentation steps may be applied, or this process can be completely skipped. Also, while the pair of eyes of an individual are natural “biological replicates” for our ArcFace-like approach, our approach may be extended to images without replicates, via current approaches for contrastive learning^14^. Furthermore, to inject labels to make more specific phenotypes, one can use a hybrid approach that minimizes both supervised and self-supervised losses.

Paradoxically, we found that the segmentation mask image, as a heavily filtered version of the raw image containing only a subset of the original information, gives more genetic association signals. They have higher heritabilities and the GWAS of which identifies most loci in the GWAS of raw image endophenotypes. We postulate that the original contrastive learning task done on the raw images was relatively easy because there are ample texture features or even artifacts to match the left and right fundus images. By replacing raw image with a vessel mask, we make the contrastive learning task harder and the model is forced to focus on vessel features.

While we prioritize the proof-of-concept, there is room for further methodological improvements. For example, it is not completely optimized to use the 128-dimensional vector as phenotypes. Moreover, the phenotyping model and segmentation model were all trained in different datasets then directly deployed to the UK Biobank data so there may exist some distribution shift that we didn’t account for. We chose not to do domain adaptation on the UK Biobank data set to avoid false association signals due to information leaking. Addressing the distribution shift may improve the separation of endophenotype distances between matched and random pairs in the UK Biobank. Also, we used the soft vessel segmentation mask instead of a binary one because the performance of the segmentation model is not good enough to permit a global threshold while at the same time preserve most details of vasculature. This might also be the cause of the appearance of many color loci in the vessel-enriched GWAS results. In addition, lack of clear image interpretation of our endophenotype derived from unsupervised learning might be a major limitation. Future work is needed to engage image interpretation methods to identify relevant image features. Moreover, our retina color GWAS uses RGB color, which may be susceptible to change in illumination. Defining retina color in other color spaces may further improve the detection power.

Admittedly, it is challenging to use an unsupervised approach to extract exclusively vessel-related information from complex modalities like retina fundus images. While our segmentation-based endophenotypes were aimed to enrich vessel-related features, they are also reflecting other related factors. One of the main factors we explored is retinal colors. Retinal color may affect the recent GWAS^8, 32, 43, 60^ on AI-based automatic extracted phenotypes from fundus images including optic nerve head morphology, retinal vessel measurements which also identified the *HERC2*/*OCA2* locus as the strongest hit. In addition, as in any association study, genetic loci identified in our study associated with retinal blood vessel could be due to secondary effect of other hidden confounding factors. For example, eye conditions such as refractive error could affect the appearance of the retina vessel. Indeed, it has been reported that *WNT7B* is associated with refractive error^61–63^. In addition, other factors such as retinal background texture were not considered. More sophisticated representation learning with disentanglement may be used to control for these correlations^64, 65^. Therefore, to establish causality relationship between the gene loci with the phenotype, further investigation, such as follow up functional experiments presented in our study, is essential.

In sum, the benefit of self-supervised-learning-derived phenotypes is that no external training labels are required. This frees up the burden of complicated and expensive labeling and makes our approach applicable to any large collection of images. As we leverage big datasets to improve our understanding of diseases, self-supervised methods are needed to efficiently extract meaningful information from medical images. We predict that iGWAS as a general phenotype discovery approach will be a fruitful research avenue.

## Methods

### Data set extraction

The DRIMDB dataset^66^, was downloaded on 2018/11/26 from https://www.researchgate.net/publication/282641760_DRIMDB_Diabetic_Retinop athy_Images_Database_Database_for_Quality_Testing_of_Retinal_Images. We used it as part of the training set to train the quality control network as it contains images with quality labels. It contains 69 bad quality fundus images and 125 good quality fundus images.

Multiple datasets (accessed in 2018/11/02), including ARIA^67^ (sample number: 143), CHASEDB1^68^ (sample number: 28), DRIVE^68, 69^ (sample number: 40), HRF^70^ (sample number: 45), IOSTAR^70, 71^ (sample number: 30) and STARE^70–73^ (sample number: 40), containing fundus images and their corresponding vessel segmentation masks, were used to train the vessel segmentation network.

The Messidor dataset^74^ (accessed in 2018/11/02) contains 1200 eye fundus color images that were acquired using a color video 3CCD camera mounted on a Topcon TRC NW6 non-mydriatic retinograph with a 45-degree field of view. The dataset was used to validate the performance of our embedding method.

The EyePACS dataset (accessed in 2018/11/02) was downloaded from Kaggle.com. It contains fundus images from both healthy subjects and subjects with different grades of diabetic retinopathy. 35,126 Kaggle training set images and 53,576 Kaggle test set images were combined. The demographic characteristics including age, sex, and ethnicity of individual images were undisclosed.

The UK Biobank data was accessed via approved project 24247. We conducted our analysis on over 65,629 British White (self-reported white British (field: 21000) and genetically identified as Caucasian (field: 22006)) participants from the UK Biobank who had fundus images available (field: 21015 and 21016). For each participant, we chose the first image for each eye, resulting in 130,329 images. Genetic data as genotyped by Applied Biosystems UK BiLEVE Axiom Array (field: 22438) and imputed (field: 22828)^75^ were downloaded. The fundus images in the UK Biobank data were taken using the TOPCON 3D OCT 1000 Mk2 alongside with the optical coherence tomography (OCT) imaging data. The data were collected in two phases: the initial assessment visit (2006-2010) at which participants were recruited and consent given and the first repeat assessment visit (2012-13). The size of each fundus image is 1536x2048 pixels.

### Image quality control

We trained a neural network to automatically assess the quality of the fundus images. Since the DRIMDB does not contain enough labelled images, we manually labeled 1,000 fundus images of good and bad quality from the EyePACS dataset and combined them with the DRIMDB dataset as the training set. An Inception v3 network^22^ pretrained on ImageNet was downloaded and fine-tuned to classify qualities of different samples with early stopping. The quality assessment network outputs a score between 0 (bad) and 1 (good) to indicate the quality of the image, and it was trained using cross entropy loss. An image was defined as good quality if the output quality score of the network from that image was greater than 0.5.

The performance of the quality assessment network was validated on a subset of UK Biobank fundus images taken from white British subjects with diabetes mellitus (n=7,683). A previously validated procedure was used to determine DM status based on self-reported DM diagnosis, use of DM medications and presence of DM complications^76^. We also used HbA1c ≥ 6.5% as a criterion for identifying DM. Two ophthalmologists were asked to grade the image for the stage of diabetic retinopathy and determine if an image is of bad quality. A fundus image in this subset was classified as bad quality if both graders agreed that the quality of the image is poor. Comparing with this ground truth, the quality assessment network reached an AUC ROC of 92.14%. At 0.5 threshold, the positive predictive value was 0.9832, the negative predictive value was 0.4916, the sensitivity was 0.7155, and the specificity was 0.9574.

### Fundus image segmentation

The graph cut algorithm^77^ from OpenCV^78^ was implemented to separate the foreground and background, and the foreground image was cropped and then resized to 224×224. The segmentation network is a specialized U-net, ESPNet^79^. ESPNet is a lightweight memory friendly architecture that allows a whole batch of images to be processed at the same time. We modified the ESPNet for the vessel segmentation task. While a typical segmentation output is a binary image mask of the original image size, we chose to use an un-binarized grey-scale image output in order to retain more details of the small vessels. The quality of the segmentation was confirmed by visual inspection. The segmentation model was trained on a combined dataset (n=306) of ARIA^67^, CHASEDB1^68^, DRIVE^68, 69^, HRF^70^, IOSTAR^70, 71^ and STARE^70–73^ for 500 epochs using the Adam optimizer^80^, with a learning rate of 1×10^-4^ using the cross-entropy loss. Then, the model with the best test Dice score^81^ was selected. On the held-out test set, the Dice score of the trained model reached 0.77.

### Embedding neural network

The raw fundus images or the retinal vessel segmentation images were fed to a network that also uses the Inception v3^22^ backbone to produce a 128-dimensional embedding vector. The final fully connected layer of the Inception v3 network was replaced to produce a 128-dimensional vector. We adopted an approach similar to ArcFace^21^: Each subject is assigned a template embedding and the network is trained to minimize the angle between embeddings of different photos of a subject and his/her template while maintaining a margin between embeddings of a specific photo and templates of different subjects. Specifically, our loss function is:

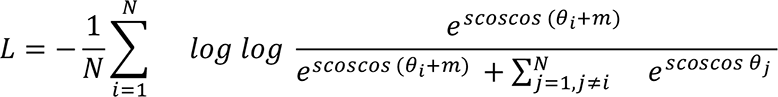

where 𝑁 is the number of samples, 𝑗 is the angle between the output of the network and the template of the 𝑗th sample, 𝑚 is the margin, and 𝑠 is the inverse temperature scaling factor. In our study, 𝑚 is set to be 30 and 𝑠 is set to be 0.5, which are the best performing hyperparameters on multiple face recognition datasets.

The embedding network was trained using 40,000 images from the EyePACS database (https://www.eyepacs.com/). The quality control network was used to score each image, and the top-ranked 40,000 images were taken. The right eye images were flipped for preprocessing, and random rotations were applied to add robustness. The training-testing split was 80/20. We also trained the embedding network with an additional task of classifying the grade of diabetic retinopathy.

The weight ratio of these two tasks was 10 to 1. The network was trained using Adam optimizer^80^ with a learning rate of 1×10^-4^ for 500 epochs on RTX 2080Ti and A100, and the model with the lowest test set loss was selected. Source code is available at https://github.com/ZhiGroup/iGWAS.

### Endophenotype GWAS

The genome-wide scans for UK Biobank were conducted over 658,720 SNPs that were directly genotyped by UK Biobank Axiom Array (field: 22438). To control for confounding factors due to ethnicity, we only included individuals of British white ethnicity (self-reported white British (field: 21000) and genetically identified as Caucasian (field: 22006)). The sample size was 65,629. We used all 130,329 images from this cohort without applying image quality control because fewer loci were identified otherwise (data not shown). The GWAS was performed with BOLT-LMM (Version 2.3.4)^31^ on all 128 dimensions of the embedding vector using the linear mixed model association method (BOLT_LMM_INF) with age, sex, and the first 10 ancestral principal components as covariates. In total, we conducted 256 GWAS, one for each of the 128 endophenotypes from one eye. As a result, each variant had 256 p-values, 128 for the left and 128 for the right fundus images. A variant was selected if the minimum of the left 128 p-values and the minimum of the right 128 p-values both passed a threshold of 5×10^-8^. The association signals from selected variants from both eyes and across endophenotypes that were merged into single independent loci if they are in linkage disequilibrium (r^2^>0.2) or within 250kb from each other.

Fine mapping was conducted for significant loci for vessel-enriched endophenotypes. For each loci group, the variant with the most significant p-value was deemed the lead SNP, and the imputed genotypes of SNPs (field: 22828) at the hit region, 250kb upstream and 250kb downstream of the lead SNP, were extracted for dense mapping. To generate consistent association results without rerunning the imputed SNPs across the entire genome, the imputed SNPs at each hit region were concatenated with the non-imputed SNPs on other chromosomes then fed into the BOLT-LMM. LocusZoom (legacy version, accessed between 2018 and 2021)^82^ was used to generate localized annotations of the significant loci groups.

### Extracting retina background color and color GWAS

The traits of retina color were created as follows. The size of each UK Biobank fundus image is 1536x2048 pixels. Right eye fundus images were first flipped before cropping. The center patch of size 400x400 pixels around the fovea region, [600:1000, 800:1200], was cropped, and the average intensities of each of 3 channels (red, green, and blue) in this patch were taken as the quantitative traits. Since the fundus images of UK Biobank are mostly aligned as they are taken with unified protocol, the patches at the same location were comparable. The 3 color variables were adjusted for in color-adjusted iGWAS. In addition, the GWAS analyses were done on the same cohorts and using the same pipeline as in vessel-enriched endophenotype GWAS.

### Genetic correlation

The genetic correlations were calculated using LDSC software (v1.0.1)^83^. Besides computing genetic correlation within the endophenotypes, we additionally selected several traits (**Supplementary Table 8, 9**) to probe the endophenotypes. We counted the number of vessel-enriched endophenotype GWAS loci that overlapped with traits from the GWAS catalog (**Supplementary Figure 7**). The selection criteria were: (1) The previous GWAS hits of the trait fall within any iGWAS loci more than twice or the traits are related to retinal or corneal disorders, and (2) The summary statistics of the trait are available from either https://alkesgroup.broadinstitute.org/UKBB/UKBB_409K/ or http://geneatlas.roslin.ed.ac.uk. To our knowledge, these are the only publicly available summary statistics computed by linear mixed models.

### Querying GWAS catalog

For each independent locus, the range from the first to the last significant SNP was first transformed using LiftOver, then a range query was performed with the range plus 250kb flanking regions on the GWAS Catalog database to identify previous associations.

### PC GWAS and heritability calculation

For vessel-enriched endophenotypes, we did PCA and selected the first 27 components (in descending order of explained variance) that explains 99% of the variance. We did GWAS on these 27 PCs on the same cohorts and using the same pipeline as in vessel-enriched endophenotype GWAS and calculated the heritabilities using LDSC^83^ v 1.0.1(https://github.com/bulik/ldsc).

### Functional validation of candidate gene involved in retina vessel development

Electroporation was performed as previously described^84^. To label retinal vessels, mice were deeply anesthetized by a sustained flow of isoflurane (3% isoflurane at 2 L/minute mixed with pure oxygen). Sulfo-NHS-LC-biotin molecules (∼226 Da, Thermo Fisher, 21335) were injected (0.5mg/g body weight) into the left ventricle of mouse hearts with a 31-gauge insulin syringe. The heartbeat was continuous for 10 minutes to ensure good circulation of the tracer. Eyes were enucleated and fixed in fresh 4% PFA for 30 minutes on ice. Retinas were dissected out in PBS, fixed overnight in 4% PFA, and washed three times in PBS the following day. Retinas were incubated with blocking buffer (containing 3% Triton X-100, 0.5% Tween 20, 1% BSA, and 10% donkey serum in PBS) at 4°C overnight. For immunohistological staining, retinas were incubated with isolectin B4 conjugated to Alexa Fluor® 647 (Molecular Probes™; I32450; 1:500) for 24 hours and washed five times in PBS. Retinas were then incubated with streptavidin conjugated to Alexa Fluor™ 488 (Thermo Fisher Scientific™; NC0186832; 1:2000) for 24 hours to detect the NHS-Biotin tracer and washed three times in PBS. Retinas were flat-mounted with Fluoromount-G® (SouthernBiotech; 0100-01) and imaged by confocal microscopy.

### Animals

Wild type mouse neonates were obtained from time pregnant CD1 mice (Charles River Laboratories, #022). All animal studies were approved by the Administrative Panel on Laboratory Animal Care (APLAC) at Stanford University.

### Data availability

The MRI and the genetic data used in this study are provided by UK Biobank (https://www.ukbiobank.ac.uk/enable-your-research/register). The summary statistics of all GWAS can be downloaded at https://drive.google.com/drive/folders/1jaQ-dCDKbY_zW0_FinPUwS7uGlNNPoMc?usp=sharing.

### Code availability

Source code is available at https://github.com/ZhiGroup/iGWAS.

## Acknowledgements

This work was supported by grants from the National Eye Institute (1R01EY032768) to Z.X., H.C., R.Chen, R Channa and D.Z., National Institute of Aging (1U01AG070112) to Z.X., W.Z., L.G., H.C., and D.Z.. In addition, this work was supported by American Diabetes Association (1-16-INI-16 to S.W. , C.L., A.D. and M. W.), NIH 1R01EY03258501 (to S.W. , C.L., A.D. and M.W.), P30 to Stanford Ophthalmology (to S.W. , C.L., A.D. and M.W.). This work was also supported by grants from the National Eye Institute (EY022356, EY018571, EY002520), Retinal Research Foundation, and NIH shared instrument grant S10OD023469 to R.Chen. R Channa is supported by the grant from the National Eye Institute (1K23EY030911-01). This work was supported in part by an Unrestricted Grant from Research to Prevent Blindness to the UW-Madison Department of Ophthalmology and Visual Sciences. L.G. was also supported by the Translational Research Institute through NASA Cooperative Agreement NNX16AO69A, NIH grants UL1TR003167 and R01NS121154, and a Cancer Prevention and Research Institute of Texas grant (RP 170668). We thank Kaitlyn Xiong for critical reading of the manuscript.

## Author contributions

R.Chen and D.Z. conceptualized the concept of iGWAS and initiated the project. Z.X., D.Z., and L.G. designed the SSuPER Model. Z.X. implemented the model, designed and conducted image analysis. R.Channa performed image grading. T.Z., Z.X. and H.C. conducted GWAS analyses. Z.X., S.K., and W.Z. conducted post-GWAS analyses. J.L., R.Chen., S.W. , C.L., A.D. and M. W. conducted experiment validation. D.Z., Z.X., R.Chen. and S.W. wrote the manuscript. All authors discussed the results and commented on the manuscript.

