## Supplementary Figures for "iGWAS: image-based genome-wide association of self-supervised deep phenotyping of human medical images"

Supplementary Information:
### Self-supervised deep phenotyping of retina fundus image reveals new genes for eye development

Ziqian Xie[1,2]
Tao Zhang[1]
Sangbae Kim[1]
Jiaxiong Lu[1]
Wanheng Zhang[3]
Cheng-Hui Lin[4]
Man-Ru Wu[4]
Alexander Davis[4]
Roomasa Channa[5]
Luca Giancardo[2]
Han Chen[3,6]
Sui Wang[4]
Rui Chen[1,*]
Degui Zhi[2,*]

[1]Department of Molecular and Human Genetics, Baylor College of Medicine, Houston, Texas 77030, USA; [2]School of Biomedical Informatics, University of Texas Health Science Center, Houston, Texas 77030, USA; [3]School of Public Health, University of Texas Health Science Center, Houston, Texas 77030, USA; [4]Department of Ophthalmology, Stanford University School of Medicine, Stanford, CA 94305, USA; [5]Department of Ophthalmology and Visual Sciences, University of Wisconsin, Madison, Wisconsin 53726; [6]Human Genetics Center, University of Texas Health Science Center, Houston, Texas 77030, USA.

Table of Content

**Supplementary Figure 1: The overall pipeline of this study.** 3

**Supplementary Figure 2: univariate distribution of 128 endophenotypes derived from both raw images (a) and vessel images (b).** 4

**Supplementary Figure 3: Genomic inflation factors of each dimension of SSuPER endophenotypes from both raw images (a) and vessel images (b). Blue: left eyes; Orange: right eyes.** 5

**Supplementary Figure 4: Scatter plot for the heritabilities of vessel-enriched endophenotypes (left) and those of raw image endophenotypes (right), directly estimated by LD score regression.** 6

**Supplementary Figure 5: LocusZoom plots for the vessel endophenotype loci. Only two loci are shown here, the rest are available at the following link: https://github.com/ZhiGroup/iGWAS/blob/master/locuszoom_plots/iGWAS/all.pdf** 6

Supplementary Figure 5.1: LocusZoom plot at rs80308281 for vasculature endophenotype GWAS. 7

Supplementary Figure 5.2: LocusZoom plot at rs9330813 for vasculature endophenotype GWAS. **Error! Bookmark not defined.**

**Supplementary Figure 6: Fundus background color Manhattan plot. The locuszoom plot for each locus is available at https://github.com/ZhiGroup/iGWAS/blob/master/locuszoom_plots/retina_color/color_all_35.pdf** 8

**Supplementary Figure 7: Phenotypes of previous GWAS hits, sorted according to the number of loci overlapping with iGWAS.** 9

**Supplementary Figure 8: Venn diagram of the number of overlapping loci between fundus background color and other traits.** 10

**References** **Error! Bookmark not defined.**

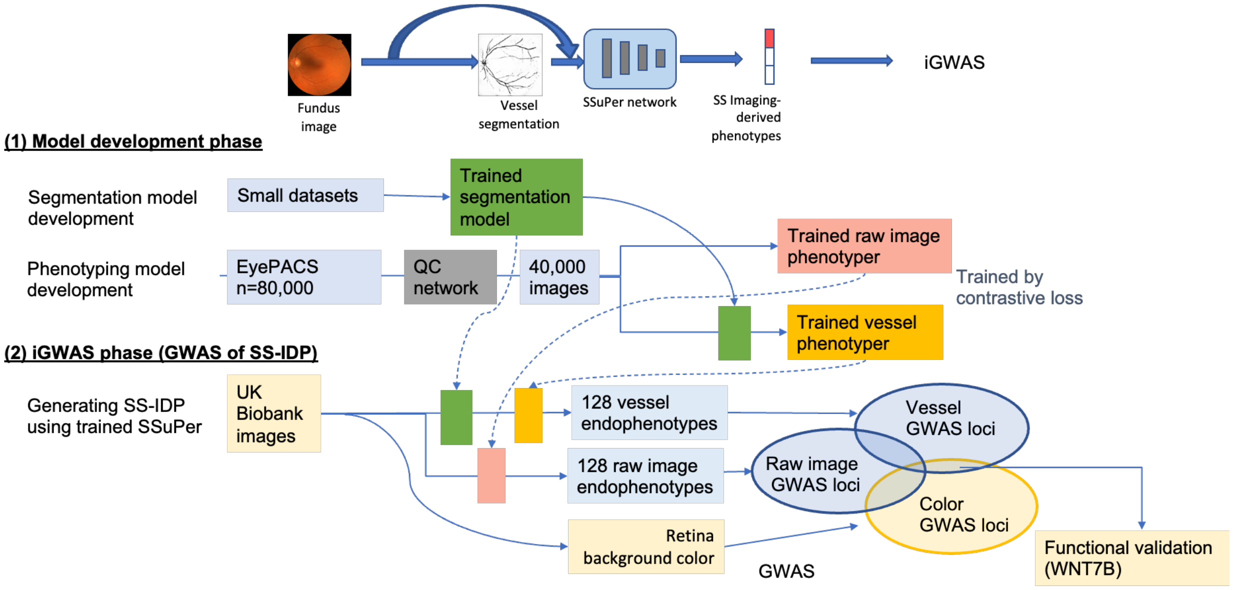


Supplementary Figure 1: The overall pipeline of this study.

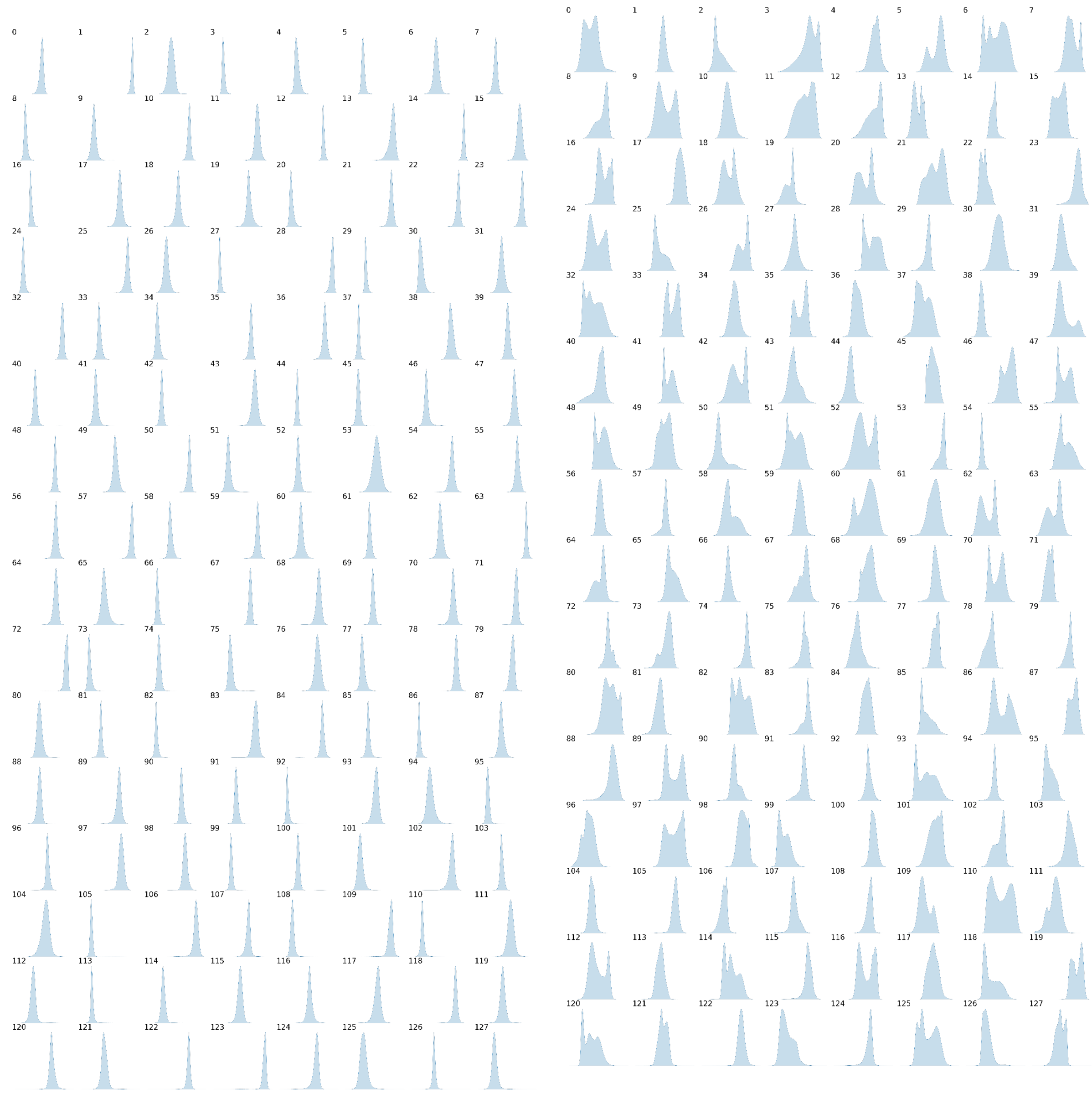

(**a**) (**b**)

Supplementary Figure 2: univariate distribution of 128 endophenotypes derived from both raw images (**a**) and vessel images (**b**).

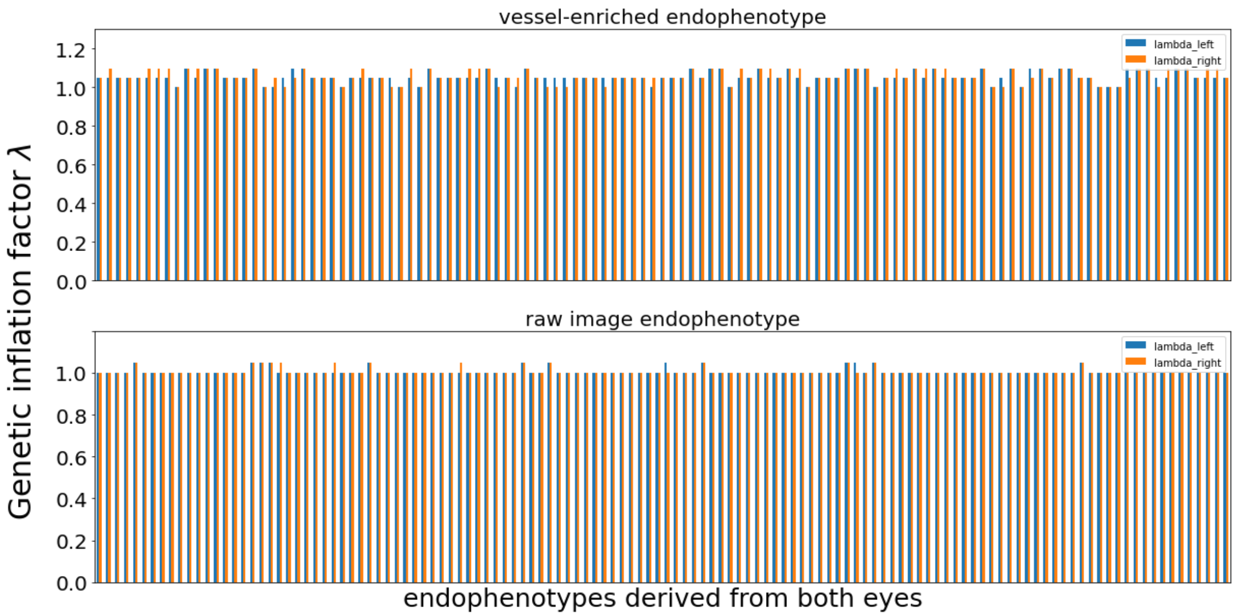


Supplementary Figure 3: Genomic inflation factors of each dimension of SSuPER endophenotypes from both raw images (**bottom**) and vessel images (**top**). Blue: left eyes; Orange: right eyes.

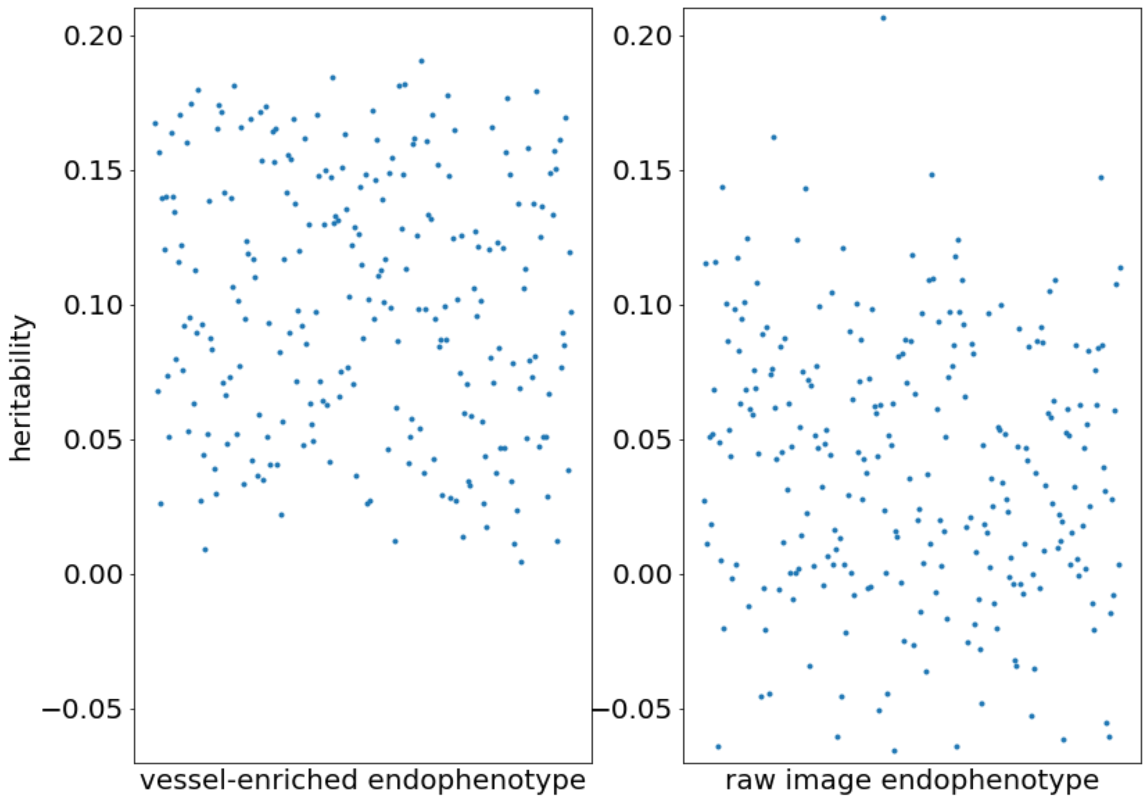


Supplementary Figure 4: Scatter plot for the heritabilities of vessel-enriched endophenotypes (left) and those of raw image endophenotypes (right), directly estimated by LD score regression.

a

b

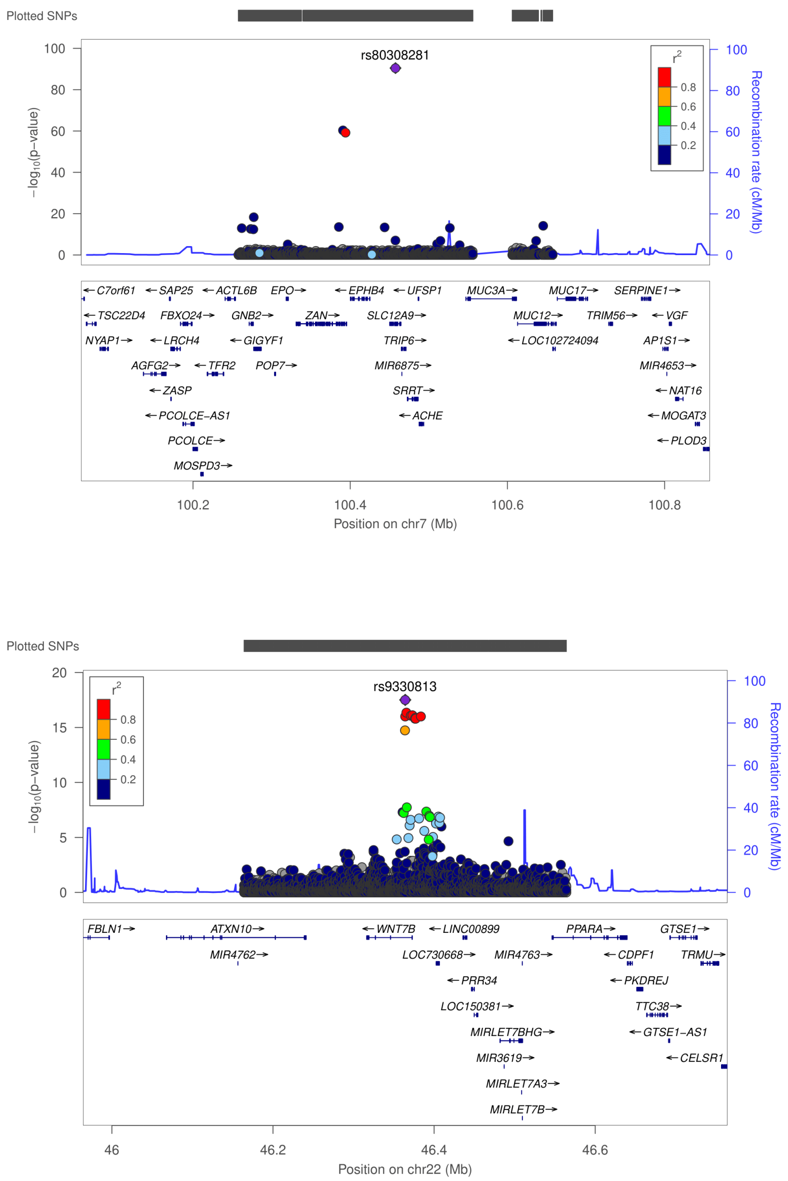


Supplementary Figure 5: LocusZoom plots for the vessel endophenotype loci (a) rs80308281; (b) rs9330813. The LocusZoom plots for remaining loci are available at the following link: https://github.com/ZhiGroup/iGWAS/blob/master/locuszoom_plots/iGWAS/all.pdf

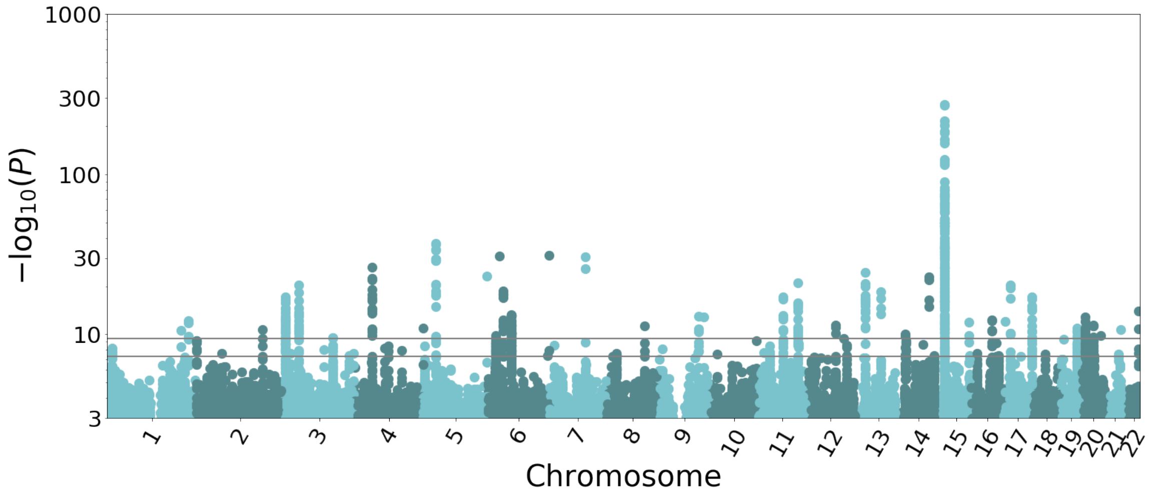


Supplementary Figure 6: Fundus background color Manhattan plot. The locuszoom plot for each locus is available at https://github.com/ZhiGroup/iGWAS/blob/master/locuszoom_plots/retina_color/color_all_35.pdf

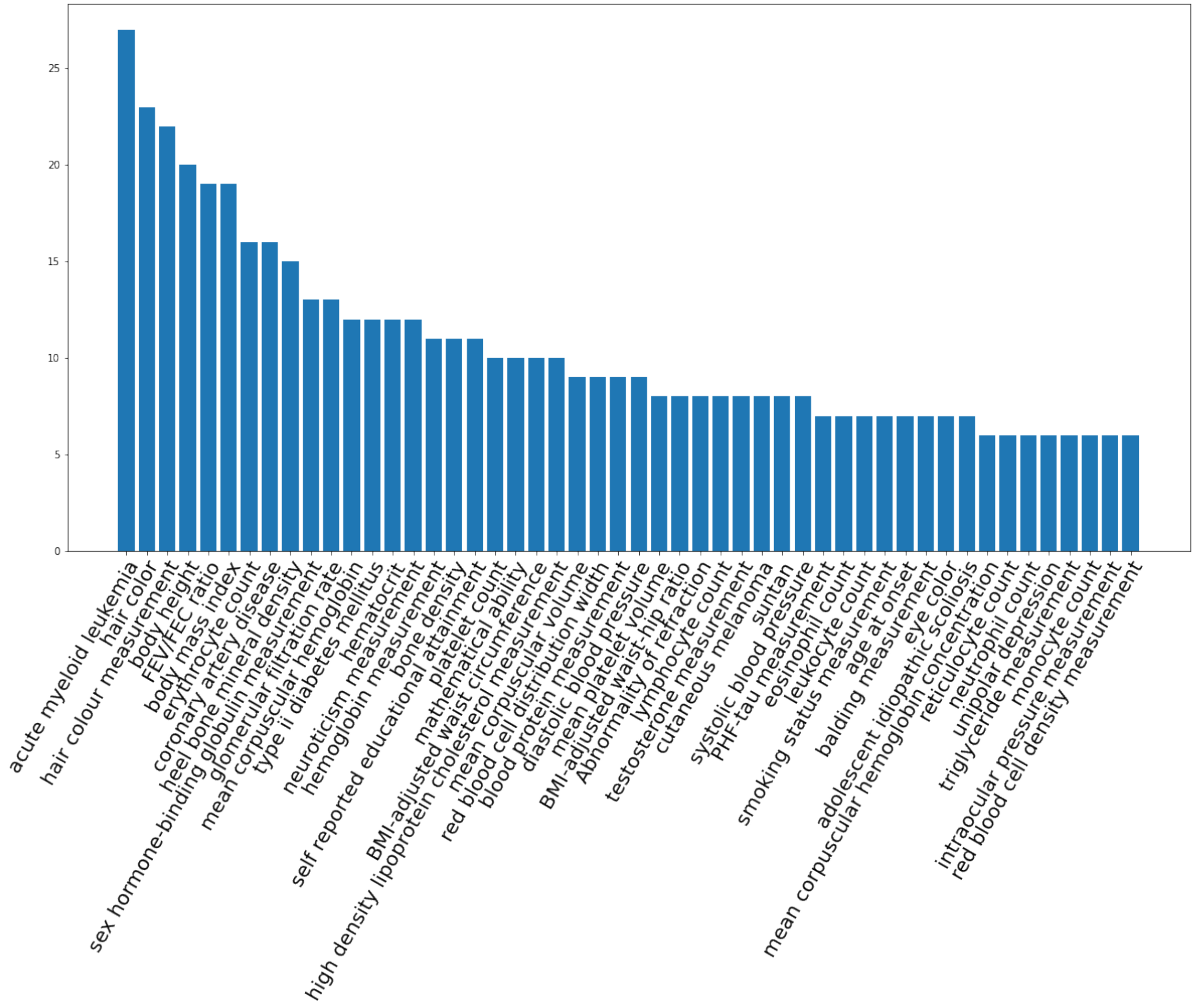


Supplementary Figure 7: Phenotypes of previous GWAS hits, sorted according to the number of loci overlapping with vessel-enriched endophenotype GWAS.

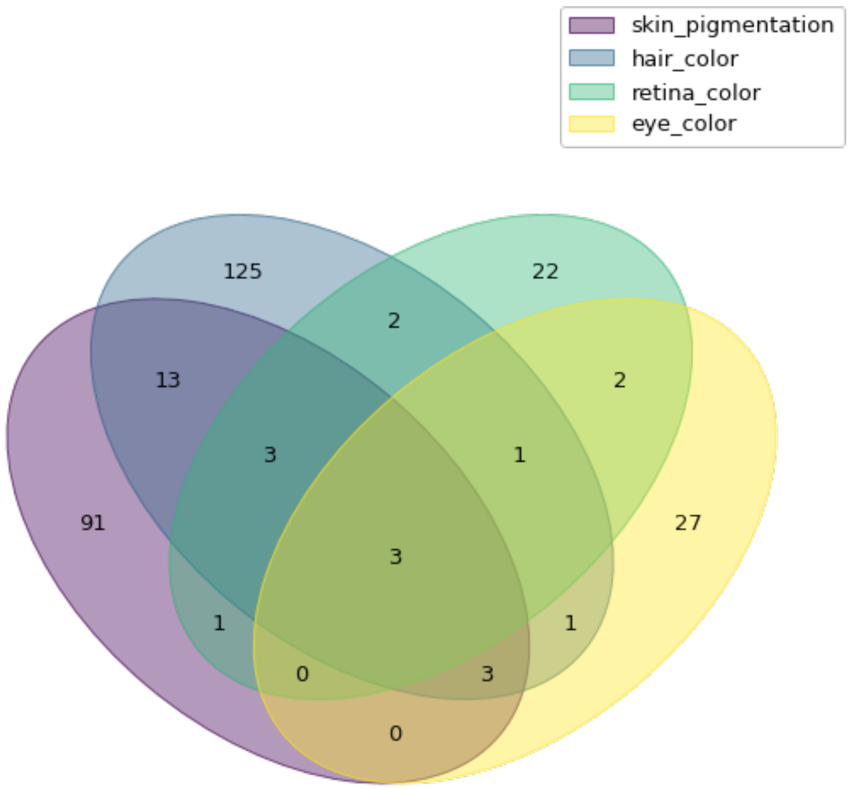


Supplementary Figure 8: Venn diagram of the number of overlapping loci between fundus background color and other traits.
